# Towards Early Warning Systems for Shigellosis in Africa: Multi-Horizon Forecasting of Incidence in Rural Western Kenya Using Statistical State-Space and Machine Learning Models

**DOI:** 10.64898/2026.10.19.26365895

**Authors:** Billy Ogwel, Brian O. Onyando, Bryan O. Nyawanda, Alex O. Awuor, Joseph Ogutu, John B. Ochieng, Maria Garcia Quesada, Patricia B. Pavlinac, Max Lau, Richard Omore, Elizabeth T Rogawski McQuade

## Abstract

**Introduction:** Despite the significant burden of Shigellosis, routine surveillance systems in resource-limited settings lack predictive capabilities to guide proactive public health responses. Consequently, we developed and evaluated the accuracy, uncertainty and computational cost of multi-horizon forecasting models for *Shigella* incidence among children <5 years in rural western Kenya using 16-year longitudinal incidence time-series.

**Methods:** We estimated monthly *Shigella* incidence using data from three pediatric enteric studies (2008– 2024), supplemented with Kenya Health Information System during inter-study gaps. Incidence estimates incorporated sampling, healthcare-seeking, stool-testing, and pathogen-specific weights, with bootstrap-derived 95% confidence intervals. Predictors included Google Trends health-related indicators, climate, temporal trends, and seasonality. Forecasts at 1-, 3-, and 6-month horizons were evaluated using rolling-origin validation. Following horizon-specific predictor selection using Boruta, 14 state-space, machine-learning, and hybrid models were compared using point-forecast and probabilistic metrics, bootstrap confidence intervals, and Model Confidence Sets. Adaptive conformal prediction recalibrated prediction intervals, while Shapley Additive exPlanations assessed predictor contributions.

**Results:** Mean monthly *Shigella* incidence was 2.51 per 100 child-years (standard deviation=1.85). *Shigella* dynamics in Kenya can be forecast beyond seasonal persistence alone for up to six months reducing absolute errors by ∼ 33%. However, optimal model architectures and predictor sets shift systematically across forecasting horizons. Temporal trend and recent incidence were the dominant predictors, while predictor importance shifted with forecast horizon toward syndromic, pharmaceutical, seasonal, and lagged environmental signals. Extreme Gradient Boosting performed best at 1 month (Root mean squared error [RMSE]=1.86), remained competitive at 3 months (RMSE=1.89), whereas exponential smoothing (ETS) performed best at 6 months (RMSE=2.05). Forecast accuracy declined with increasing horizon. ETS and TBATS provided the best-calibrated prediction intervals, whereas machine-learning models generally under-covered. Model Confidence Sets demonstrated horizon-dependent model distinguishability.

**Conclusion:** Horizon-specific model selection balancing predictive accuracy, uncertainty calibration, and computational feasibility could strengthen operational early warning systems for shigellosis.

**Author Summary:** *Shigella* infections remain an important cause of illness among young children in low-resource settings, yet public health systems often lack tools to anticipate changes in disease burden before they occur. Early warning systems that can reliably predict future *Shigella* incidence could help health authorities plan surveillance, allocate resources, and prepare for periods of increased transmission. We used 16 years of data from children <5 years in rural western Kenya to develop and compare different approaches for forecasting monthly *Shigella* incidence 1-, 3-, and 6-months ahead. We combined disease surveillance data with information on seasonal patterns, weather, and health-related internet searches. We found that *Shigella* incidence could be predicted beyond what would be expected from seasonal patterns alone, with useful forecasts possible up to six months ahead. However, the best forecasting approach depended on how far into the future predictions were required. Machine-learning methods provided the most accurate short-term predictions, while simpler statistical models performed better for longer-term forecasts and provided more reliable estimates of uncertainty. These findings highlight the importance of selecting forecasting methods according to the intended prediction horizon and operational requirements. Such approaches could strengthen early warning and preparedness for childhood *Shigella* infections in resource-limited settings.

## Introduction

*Shigella* remains a leading cause of diarrheal morbidity and mortality in young children, with recent World Health Organization (WHO) estimates for 2021 attributing approximately 33 million cases of diarrheal disease and 50,164 deaths among children under five years of age [1]. This burden is disproportionately concentrated in sub-Saharan Africa and South Asia, where inadequate water, sanitation, and hygiene (WASH) infrastructure, high population density, malnutrition, and limited access to healthcare continue to facilitate transmission. More recent estimates from the Enterics for Global Health (EFGH) study, conducted across 7 countries in Africa, Asia and Latin America reported an overall *Shigella* incidence of 6.4 cases per 100 child-years based on culture and 14.6 cases per 100 child-years based on quantitative PCR (qPCR) among children aged 6–35 [2]. In Kenya, the corresponding incidence estimates were 5.5 and 11.2 cases per 100 child-years. Shigellosis also imposes a substantial economic burden, with the average cost per episode in EFGH estimated at approximately US$9 and US$44 for outpatient and inpatient care, respectively, across all study sites, compared with US$10 and US$45 per episode in Kenya [3]. Beyond acute illness, *Shigella* infection has been associated with adverse long-term outcomes, including prolonged diarrhea, growth faltering, impaired cognitive development, and increased susceptibility to subsequent infections [4].

Despite this significant burden, routine surveillance systems in resource-limited settings, including in Kenya, lack predictive capabilities to guide proactive public health responses. Unlike cholera and typhoid, which benefit from established early warning systems [5,6], *Shigella* surveillance remains largely passive, relying on clinical syndromic reporting from peripheral health facilities without laboratory confirmation. This reactive approach fails to capture the true incidence trajectory, as mild cases may fail to seek care, while severe cases receive empirical treatment without diagnostic confirmation, further driving antimicrobial resistance [7–9]. Forecasting is particularly relevant for *Shigella* given increasing concerns regarding antimicrobial resistance, and the need to optimize surveillance systems in resource-constrained settings. Anticipating periods of increased disease activity could help health systems strengthen preparedness and improve response strategies before transmission peaks occur. Waterborne and foodborne enteric pathogens often exhibit predictable temporal dynamics driven by rainfall, temperature, and land use patterns [10,11]. Western Kenya experiences a bimodal rainfall pattern with long rains occurring from March to May and short rains from October to December [12], each associated with increased surface runoff contaminating shallow wells and riverine water sources. These recurring environmental conditions may influence *Shigella* transmission dynamics, creating opportunities to anticipate periods of elevated disease risk and support the development of forecasting models for early warning and public health preparedness.

Existing literature on enteric disease forecasting has prioritized cholera, typhoid, and cryptosporidiosis, with *Shigella* receiving comparatively limited attention, particularly in sub-Saharan Africa [13]. Existing *Shigella* forecasting studies have largely been conducted in Asia [14–16] and have often evaluated a limited range of modeling approaches. Evidence is especially limited from African settings, where disease transmission patterns, environmental drivers, healthcare-seeking behavior, and surveillance systems may differ substantially from those observed elsewhere. As a result, there remains uncertainty regarding which forecasting approaches provide the most accurate and operationally useful predictions for *Shigella* incidence in endemic African populations. Consequently, we assembled a 16-year longitudinal incidence time series by integrating data from three pediatric enteric disease studies conducted in Siaya County (2008–2024) and supplementing inter-study gaps with Kenya Health Information System (KHIS) surveillance data. We developed and evaluated the accuracy, uncertainty and computational cost of time-series forecasting models for *Shigella* incidence among children under five years of age in rural western Kenya across three horizons (1, 3 and 6 months) using this harmonized dataset.

## Results

### Shigella Incidence

Over the 16-year period, the overall mean monthly *Shigella* MSD incidence (standard deviation [±SD]) was 2.51 per 100 child-years (±1.85) (Fig 1). By individual study, the mean (±SD) incidences were 2.05 (±1.32), 0.56 (±0.26), 2.13 (±1.68), and 3.02 (±1.37) per 100 child-years for GEMS, GEMS_1a, VIDA and EFGH, respectively. Inter-study gaps filled using KHIS data exhibited a mean incidence of 3.65 ± (2.0).

**Fig 1.**
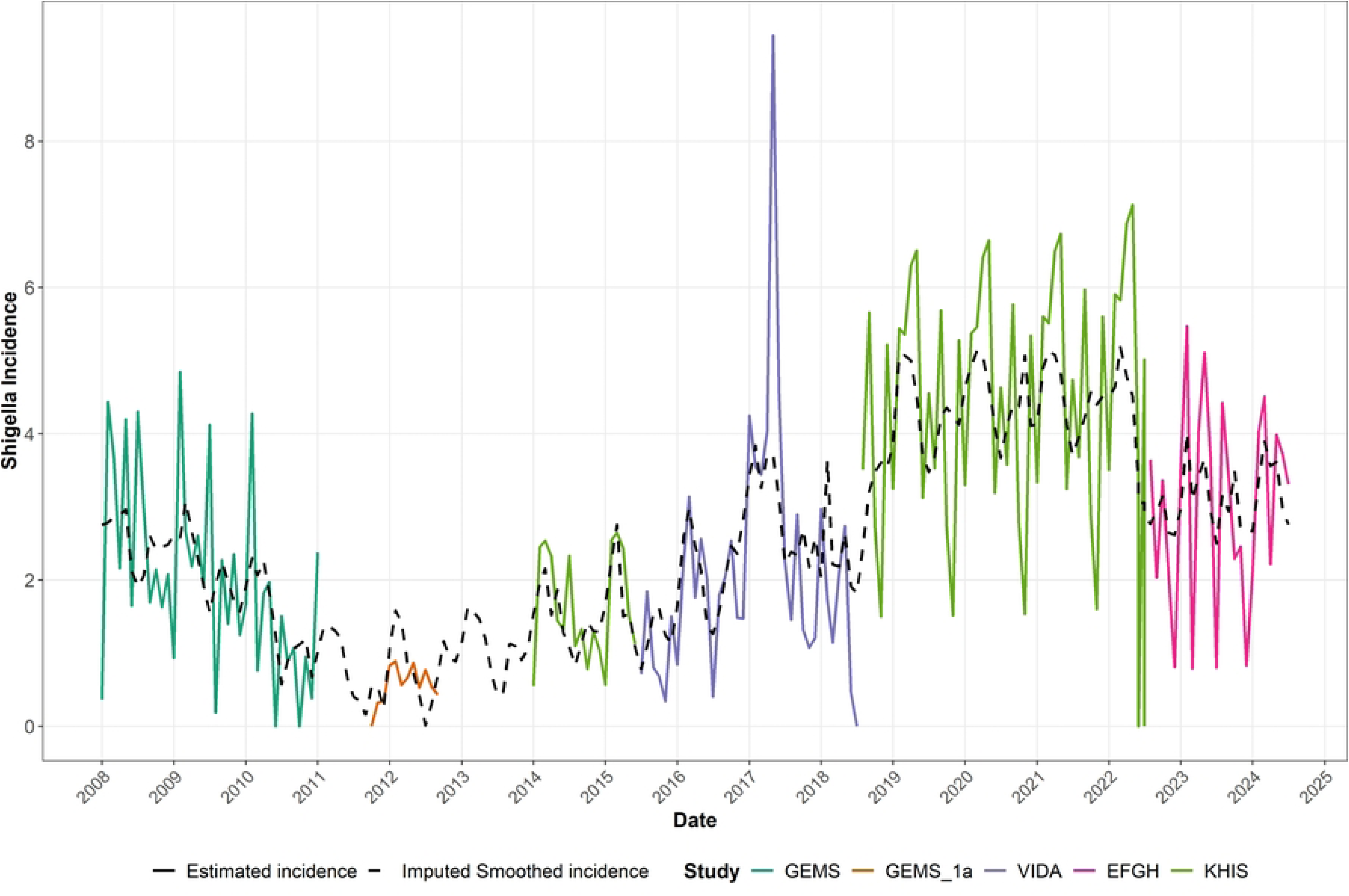
Monthly *Shigella* incidence estimates among children aged <5 years in Siaya County, Kenya, derived from integrated surveillance data, 2008–2024

### Predictor Selection and Importance Across Horizons

Time-aware Boruta analysis identified temporal and autoregressive predictors as the dominant predictors, with their relative contribution changing across forecast horizons (Fig 2 and S1 Fig). The linear trend (t) had the highest mean importance overall (0.32; h=1: 0.33, h=3: 0.33, h=6: 0.29) and was selected in 47.9% of windows, while Historical incidence (1-month incidence lag) was the most consistently selected predictor (54.2%) and ranked second in importance (0.20), declining from 0.23 at h=1 to 0.16 at h=6. 2-month incidence lag and 3-month incidence lag also showed decreasing importance (0.16 and 0.13, respectively).

**Fig 2.**
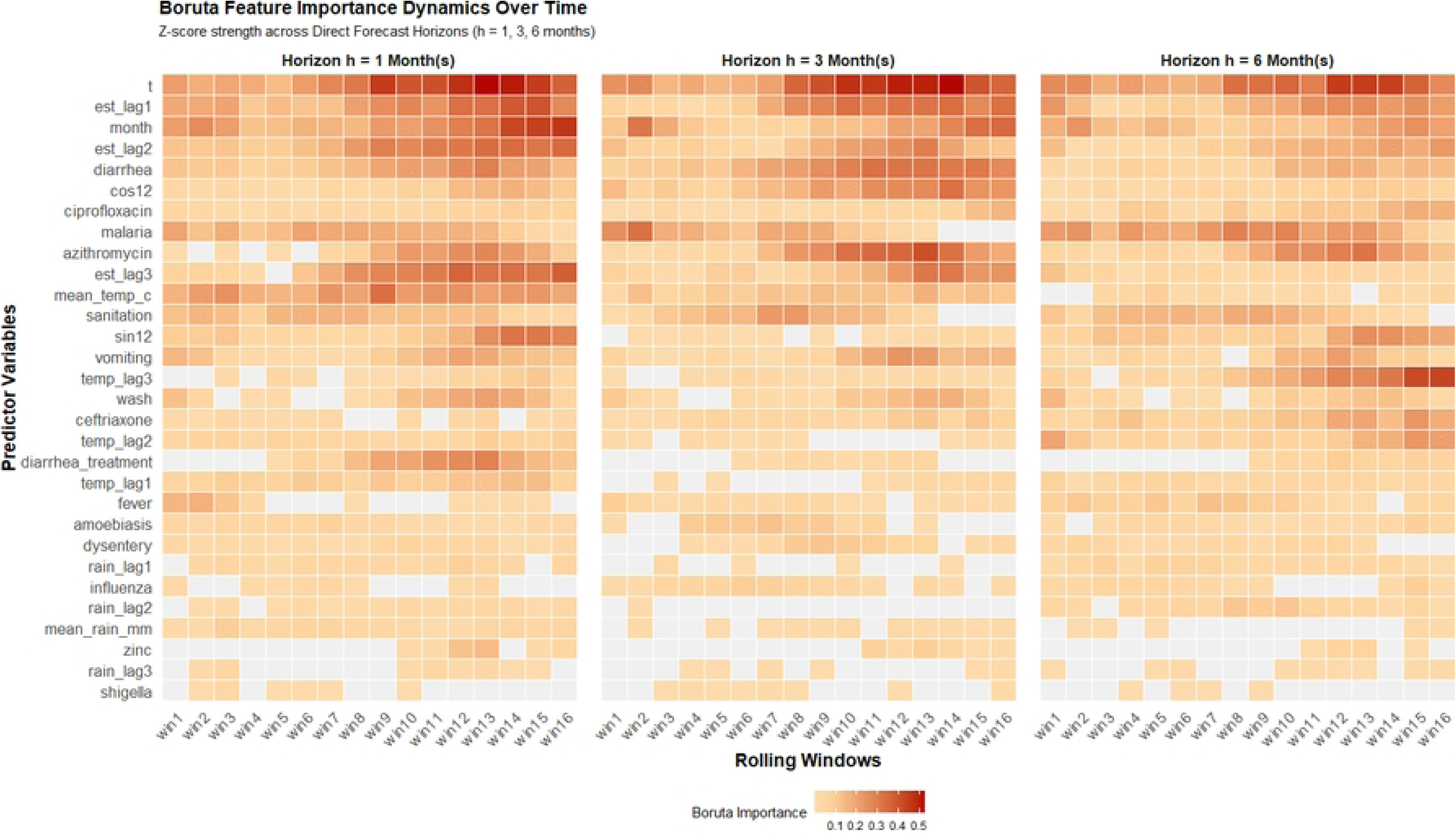
Boruta-derived predictor importance across forecasting horizons for *Shigella* incidence, 2008– 2024.

At h=1, recent incidence and seasonality dominated: month and 3-month incidence lag were selected in 62.5% of windows, 1-month incidence lag in 56.2%, and concurrent temperature in 50.0%; While 1-month incidence lag (0.33), month (0.25), and concurrent temperature (0.22) had the highest mean importance.

At h=3, prediction shifted toward syndromic and pharmaceutical signals, with 1-month incidence lag, diarrhea, and t each selected in 62.5% of windows and azithromycin in 50.0%. The leading predictors in terms of mean importance were diarrhea (0.22), azithromycin (0.19), and cosine harmonic transform (cos12) (0.18). At h=6, short-term autoregressive effects weakened (3-month incidence lag was never selected), while t, month, 1-month incidence lag, and 2-month incidence lag remained selected in 31.2%, 43.8%, 43.8%, and 37.5% of windows, respectively. Lagged temperature emerged as more important, with 3-month lag selected in 37.5% of windows and mean importance increasing to 0.17, whereas concurrent temperature declined to 0.03. Malaria also increased in importance from 0.14 at h=1 to 0.20 at h=6, potentially reflecting shared temporal or seasonal dynamics rather than causality (Figure 2 and S1 Fig).

Across domains, autoregressive predictors had the greatest selection stability, declining from 56.2% at h=1 to 27.1% at h=6, while temporal/seasonal predictors remained comparatively stable (35.9%, 29.7%, and 25.0%). Clinical and pharmaceutical predictors, particularly diarrhea and azithromycin, provided complementary horizon-specific information. In contrast, rainfall contributed little (mean importance 0.01; 3-month lag: −0.01), as did *Shigella* search volume (−0.01), while ciprofloxacin, ceftriaxone, wash, and vomiting had modest importance (Figure 2 and S1 Fig). Overall, forecasting was primarily anchored by temporal trends and recent incidence, supplemented by seasonal, syndromic, pharmaceutical, and lagged environmental signals whose relevance varied systematically with forecast horizon. These findings reflect predictive contribution rather than causality.

### Deterministic Model Performance

Evaluation of the forecasting architectures across the three temporal horizons revealed a distinct transition in optimal model archetypes as the forecasting window extended. Overall, the best-performing models consistently outperformed the seasonal-naïve benchmark across all forecast horizons, demonstrating added predictive value beyond simply extrapolating seasonal recurrence. Nevertheless, absolute predictive error increased with forecast horizon, indicating diminishing forecastability of *Shigella*incidence over longer prediction windows.

The hybrid models generally did not provide additional gains over the strongest individual models. More specifically, at the 1-month horizon, machine-learning models provided the strongest overall performance, with XGBoost achieving the lowest RMSE (1.86, 95% CI: 1.57–2.15), NRMSE (0.96, 0.85–1.06), MAE (1.37, 1.19–1.55), MASE (1.45, 1.24–1.70), and sMAPE (47.27%, 41.08–53.66), as well as the highest Pearson correlation (r=0.40, 0.25–0.56). RF showed similar performance (RMSE 1.89, 1.63–2.19; r=0.35, 0.19–0.50). Among conventional time-series models, ETS performed best, with RMSE 1.95 (1.69–2.21), whereas the performance of Prophet and NBGLM were suboptimal. The hybrid models did not improve consistently over their constituent models. Compared with the seasonal-naïve benchmark (RMSE 2.80, MAE 1.75, MASE 1.87, sMAPE 52.29%, r=0.29), the best-performing model, XGBoost, reduced RMSE by 33.6%, MAE by 21.7%, MASE by 22.5%, and sMAPE by 9.6% (Table 1).

**Table 1:** Deterministic model performance across horizons in *Shigella* forecasting, 2008-2024.

| Horizon | Algorithm | RMSE [95% CI] | NRMSE [95% CI] | MAE [95% CI] | MASE [95% CI] | sMAPE [95% CI] | Pearson_R [95% CI] |
| --- | --- | --- | --- | --- | --- | --- | --- |
| 1 | ARIMAX | 2.09 [1.85 – 2.34] | 1.08 [0.99 – 1.17] | 1.62 [1.44 – 1.82] | 1.66 [1.45 – 1.89] | 52.08 [45.95 – 58.80] | 0.24 [0.10 – 0.39] |
|  | TBATS | 2.02 [1.77 – 2.27] | 1.04 [0.95 – 1.13] | 1.60 [1.43 – 1.78] | 1.70 [1.50 – 1.94] | 53.89 [47.76 – 60.34] | 0.28 [0.14 – 0.42] |
|  | ETS | 1.95 [1.69 – 2.21] | 1.01 [0.92 – 1.10] | 1.54 [1.37 – 1.71] | 1.61 [1.42 – 1.84] | 50.31 [44.62 – 56.55] | 0.30 [0.16 – 0.45] |
|  | Prophet | 3.67 [2.38 – 4.84] | 1.90 [1.23 – 2.49] | 2.02 [1.60 – 2.49] | 2.04 [1.63 – 2.48] | 53.20 [46.04 – 60.38] | 0.11 [-0.02 – 0.30] |
|  | NBGLM | 2.83 [2.34 – 3.31] | 1.46 [1.22 – 1.76] | 2.02 [1.75 – 2.29] | 2.14 [1.83 – 2.44] | 56.29 [50.53 – 63.06] | 0.17 [0.03 – 0.31] |
|  | GAM | 2.06 [1.84 – 2.28] | 1.07 [0.98 – 1.16] | 1.59 [1.40 – 1.77] | 1.65 [1.45 – 1.86] | 51.10 [44.71 – 57.35] | 0.28 [0.15 – 0.40] |
|  | RF | 1.89 [1.63 – 2.19] | 0.98 [0.88 – 1.07] | 1.42 [1.25 – 1.61] | 1.50 [1.29 – 1.75] | 48.24 [42.39 – 54.87] | 0.35 [0.19 – 0.50] |
|  | SVR | 2.36 [2.08 – 2.64] | 1.22 [1.10 – 1.34] | 1.78 [1.57 – 2.00] | 1.83 [1.61 – 2.10] | 57.11 [50.82 – 64.11] | 0.25 [0.09 – 0.39] |
|  | **XGBoost** | **1.86 [1.57 – 2.15]** | **0.96 [0.85 – 1.06]** | **1.37 [1.19 – 1.55]** | **1.45 [1.24 – 1.70]** | **47.27 [41.08 – 53.66]** | **0.40 [0.25 – 0.56]** |
|  | HYBRID_ARIMAX_XGB | 2.10 [1.78 – 2.36] | 1.13 [0.99 – 1.28] | 1.61 [1.39 – 1.83] | 1.47 [1.24 – 1.69] | 50.00 [42.14 – 57.89] | 0.29 [0.13 – 0.45] |
|  | HYBRID_TBATS_XGB | 2.10 [1.79 – 2.41] | 1.09 [0.96 – 1.20] | 1.57 [1.38 – 1.77] | 1.67 [1.45 – 1.92] | 58.19 [50.22 – 65.71] | 0.38 [0.23 – 0.53] |
|  | HYBRID_ARIMAX_RF | 2.02 [1.80 – 2.24] | 1.10 [0.98 – 1.21] | 1.63 [1.45 – 1.81] | 1.46 [1.28 – 1.65] | 49.73 [42.96 – 56.79] | 0.32 [0.18 – 0.46] |
|  | HYBRID_TBATS_RF | 1.97 [1.68 – 2.26] | 1.02 [0.91 – 1.12] | 1.48 [1.30 – 1.67] | 1.59 [1.38 – 1.83] | 51.55 [45.31 – 58.42] | 0.38 [0.23 – 0.52] |
|  | SEASONAL_NAIVE | 2.80 [0.91 – 3.99] | 1.02 [0.45 – 1.47] | 1.75 [0.81 – 2.84] | 1.87 [0.81 – 3.31] | 52.29 [31.24 – 75.55] | 0.29 [-0.25 – 0.93] |
| 3 | ARIMAX | 2.57 [2.23 – 2.91] | 1.37 [1.22 – 1.52] | 2.03 [1.80 – 2.28] | 2.06 [1.80 – 2.33] | 58.96 [51.55 – 66.00] | 0.03 [-0.15 – 0.21] |
|  | TBATS | 2.04 [1.76 – 2.32] | 1.09 [0.97 – 1.20] | 1.59 [1.39 – 1.80] | 1.68 [1.43 – 1.93] | 53.37 [46.09 – 60.77] | 0.21 [0.06 – 0.39] |
|  | ETS | 1.93 [1.66 – 2.20] | 1.03 [0.91 – 1.13] | 1.52 [1.34 – 1.71] | 1.60 [1.37 – 1.85] | 50.38 [43.63 – 57.59] | 0.28 [0.12 – 0.46] |
|  | Prophet | 6.08 [3.68 – 7.84] | 3.24 [1.98 – 4.25] | 2.82 [1.98 – 3.71] | 2.72 [2.04 – 3.44] | 61.23 [52.46 – 70.50] | 0.03 [-0.11 – 0.23] |
|  | NBGLM | 4.08 [3.31 – 4.86] | 2.17 [1.79 – 2.60] | 2.84 [2.43 – 3.29] | 2.93 [2.47 – 3.45] | 67.75 [60.12 – 75.02] | -0.01 [-0.20 – 0.18] |
|  | GAM | 2.16 [1.87 – 2.43] | 1.15 [1.03 – 1.27] | 1.71 [1.51 – 1.90] | 1.76 [1.54 – 1.99] | 54.20 [46.36 – 61.47] | 0.22 [0.05 – 0.39] |
|  | RF | 1.89 [1.63 – 2.18] | 1.01 [0.91 – 1.10] | 1.45 [1.27 – 1.65] | 1.53 [1.31 – 1.77] | 48.05 [41.08 – 56.04] | 0.28 [0.10 – 0.45] |
|  | SVR | 2.44 [2.11 – 2.73] | 1.30 [1.15 – 1.46] | 1.87 [1.63 – 2.10] | 1.87 [1.64 – 2.15] | 58.06 [51.40 – 66.12] | 0.10 [-0.06 – 0.28] |
|  | **XGBoost** | **1.89 [1.61 – 2.20]** | **1.01 [0.89 – 1.12]** | **1.43 [1.23 – 1.63]** | **1.49 [1.26 – 1.74]** | **48.88 [42.14 – 56.83]** | **0.35 [0.18 – 0.53]** |
|  | HYBRID_ARIMAX_XGBoost | 2.50 [1.93 – 2.98] | 1.34 [1.15 – 1.56] | 1.75 [1.35 – 2.10] | 1.63 [1.25 – 2.04] | 46.84 [36.51 – 56.79] | 0.06 [-0.18 – 0.33] |
|  | HYBRID_TBATS_XGBoost | 2.02 [1.73 – 2.29] | 1.07 [0.94 – 1.19] | 1.52 [1.31 – 1.74] | 1.61 [1.37 – 1.89] | 52.50 [45.26 – 60.39] | 0.35 [0.19 – 0.50] |
|  | HYBRID_ARIMAX_RF | 2.60 [2.07 – 3.06] | 1.39 [1.21 – 1.60] | 1.94 [1.58 – 2.28] | 1.78 [1.44 – 2.17] | 51.57 [41.83 – 61.80] | 0.00 [-0.23 – 0.27] |
|  | HYBRID_TBATS_RF | 2.00 [1.73 – 2.27] | 1.07 [0.95 – 1.18] | 1.53 [1.32 – 1.74] | 1.63 [1.38 – 1.90] | 51.09 [43.46 – 58.63] | 0.30 [0.15 – 0.46] |
|  | SEASONAL_NAIVE | 2.66 [2.18 – 3.09] | 1.45 [1.22 – 1.77] | 2.12 [1.71 – 2.57] | 2.16 [1.71 – 2.63] | 70.57 [55.41 – 86.24] | 0.11 [-0.12 – 0.34] |
| 6 | ARIMAX | 2.30 [1.97 – 2.65] | 1.21 [1.09 – 1.36] | 1.88 [1.63 – 2.14] | 1.99 [1.65 – 2.33] | 57.84 [48.84 – 65.93] | 0.13 [-0.08 – 0.33] |
|  | TBATS | 2.18 [1.83 – 2.57] | 1.14 [1.02 – 1.28] | 1.73 [1.47 – 2.00] | 1.83 [1.50 – 2.17] | 53.38 [44.64 – 61.74] | 0.15 [-0.06 – 0.35] |
|  | **ETS** | **2.05 [1.71 – 2.43]** | **1.07 [0.96 – 1.20]** | **1.63 [1.40 – 1.87]** | **1.73 [1.42 – 2.07]** | **51.16 [42.82 – 59.48]** | **0.20 [-0.00 – 0.39]** |
|  | Prophet | 5.84 [3.85 – 7.55] | 3.06 [2.03 – 4.16] | 3.24 [2.33 – 4.25] | 3.14 [2.31 – 3.97] | 71.79 [59.52 – 83.65] | 0.00 [-0.16 – 0.20] |
|  | NBGLM | 4.28 [2.77 – 5.87] | 2.24 [1.49 – 3.12] | 2.72 [2.13 – 3.42] | 2.84 [2.22 – 3.61] | 66.67 [58.19 – 75.15] | 0.21 [0.01 – 0.38] |
|  | GAM | 2.66 [2.26 – 3.09] | 1.39 [1.22 – 1.65] | 2.11 [1.85 – 2.41] | 2.20 [1.89 – 2.58] | 63.49 [55.10 – 71.99] | 0.19 [0.00 – 0.39] |
|  | RF | 2.14 [1.81 – 2.48] | 1.12 [1.01 – 1.25] | 1.73 [1.49 – 1.98] | 1.84 [1.53 – 2.18] | 57.00 [48.64 – 65.33] | 0.16 [-0.04 – 0.37] |
|  | SVR | 2.40 [2.06 – 2.79] | 1.26 [1.12 – 1.42] | 1.91 [1.63 – 2.19] | 1.98 [1.64 – 2.34] | 65.29 [56.71 – 72.99] | 0.26 [0.07 – 0.45] |
|  | XGBoost | 2.17 [1.83 – 2.49] | 1.14 [1.00 – 1.28] | 1.74 [1.49 – 1.99] | 1.83 [1.51 – 2.18] | 57.76 [48.99 – 66.53] | 0.21 [0.03 – 0.41] |
|  | HYBRID_ARIMAX_XGBoost | 2.28 [1.84 – 2.70] | 1.23 [1.04 – 1.47] | 1.78 [1.44 – 2.10] | 1.80 [1.38 – 2.26] | 56.15 [45.29 – 64.93] | 0.27 [0.04 – 0.52] |
|  | HYBRID_TBATS_XGBoost | 2.24 [1.94 – 2.62] | 1.18 [1.04 – 1.35] | 1.80 [1.54 – 2.10] | 1.87 [1.55 – 2.21] | 58.87 [49.22 – 68.86] | 0.25 [0.06 – 0.42] |
|  | HYBRID_ARIMAX_RF | 2.44 [1.96 – 2.88] | 1.32 [1.14 – 1.59] | 1.94 [1.60 – 2.27] | 1.95 [1.54 – 2.41] | 59.90 [49.09 – 69.09] | 0.19 [-0.04 – 0.42] |
|  | HYBRID_TBATS_RF | 2.26 [1.96 – 2.63] | 1.19 [1.06 – 1.34] | 1.83 [1.57 – 2.10] | 1.92 [1.60 – 2.27] | 57.82 [49.52 – 66.72] | 0.20 [-0.01 – 0.38] |
|  | SEASONAL_NAIVE | 2.94 [2.52 – 3.38] | 1.54 [1.39 – 1.75] | 2.34 [1.99 – 2.68] | 2.37 [1.99 – 2.79] | 74.04 [62.86 – 84.69] | -0.13 [-0.29 – 0.07] |

\*RF-Random Forest; XGBOOST- Extreme Gradient Boosting; SVR- Support Vector Regression; GAM-General Additive Models; NBGLM-Negative Binomial Generalized Linear Models; RMSE-Root Mean Square Error; NRMSE-Normalized- Root Mean Square Error MAE-Mean Absolute Error; MASE-Mean Absolute Scaled Error sMAPE-Symmetric Mean Absolute Percentage Error

At the 3-month horizon, performance generally deteriorated relative to h=1, particularly for ARIMAX, Prophet, and NBGLM. RF and XGBoost remained the strongest models, both achieving RMSE of 1.89 (95%CI, RF: 1.63–2.18; XGBoost: 1.61–2.20), although XGBoost had a lower MAE at 1.43 (1.23–1.63), while RF had 1.45 (1.27–1.65). XGBoost also showed the highest Pearson correlation (r=0.35, 0.18–0.53). ETS remained the strongest conventional model (RMSE 1.93, 1.66–2.20; MAE 1.52, 1.34–1.71). The ARIMAX-XGB hybrid had a lower sMAPE (46.84%, 36.51–56.79) than the individual models but did not achieve comparable RMSE or correlation performance. Relative to seasonal naïve (RMSE 2.66, MAE 2.12, MASE 2.16, sMAPE 70.57%, r=0.11), XGBoost reduced RMSE by 29.0%, MAE by 32.5%, MASE by 31.0%, and sMAPE by 30.7%, while providing a higher correlation with observed incidence (Table 1).

At the 6-month horizon, predictive errors increased further and the relative advantage of machine-learning models diminished. ETS provided the lowest RMSE (2.05, 1.71–2.43), NRMSE (1.07, 0.96–1.20), and MAE (1.63, 1.40–1.87), while RF and XGBoost remained competitive (RMSE 2.14 and 2.17, respectively). ETS also had the lowest sMAPE among the principal models (51.16%, 42.82–59.48). Pearson correlations were generally weaker at this horizon, with SVR showing the highest point estimate (r=0.26, 0.07–0.45), although its error metrics were comparatively poorer. Prophet, NBGLM, GAM, and the seasonal-naïve model showed substantially poorer predictive accuracy at longer horizons. Compared with seasonal naïve (RMSE 2.94, MAE 2.34, MASE 2.37, sMAPE 74.04%, r=−0.13), ETS reduced RMSE by 30.3%, MAE by 30.3%, MASE by 27.0%, and sMAPE by 30.9%; its Pearson correlation was also higher (r=0.20, −0.00–0.39) (Table 1). The decline in accuracy at longer horizons is expected due to increasing uncertainty and non-stationarity in the incidence series (Table 1).

### Calibration and Probabilistic Uncertainty

Calibration and probabilistic forecast performance varied substantially across models and horizons. ETS provided the most consistently well-calibrated forecasts, with coverage of 0.88 at both 1-and 3-month horizons and 0.80 at 6 months, while also achieving the lowest Winkler scores (11.50, 11.20, and 12.99) and Pinball losses (0.14, 0.14, and 0.16), respectively. TBATS showed a similar pattern, with coverage of 0.85–0.82–0.77 and relatively low Winkler and Pinball scores across the three horizons. These models therefore provided a favorable balance between interval coverage and sharpness (Table 2). In contrast, XGBoost achieved relatively high hit rates (0.72, 0.71, and 0.62) but substantially under-covered, with coverage ranging from 0.19 at 1 month to 0.29 at 3 months and 0.33 at 6 months. Its relatively high Winkler (40.12–40.41–39.04) and Pinball scores (0.50–0.51–0.49) further indicated poorly calibrated probabilistic forecasts. The hybrid XGBoost models showed similarly low coverage, particularly at longer horizons, suggesting that combining these models did not improve uncertainty calibration. The seasonal-naive benchmark generally achieved better coverage than most machine-learning models, particularly at 3 and 6 months (0.73 and 0.75), although its uncertainty estimates were less sharp than ETS/TBATS. At 1 month, seasonal naive had very high hit rate (0.93) and coverage (0.81), but with wide confidence intervals reflecting greater uncertainty. Overall, ETS and TBATS provided the strongest probabilistic forecasts, maintaining comparatively high coverage with low Winkler, Pinball, and CRPS scores, whereas several ML and hybrid models produced narrow prediction intervals with under-coverage (Table 2).

**Table 2.**
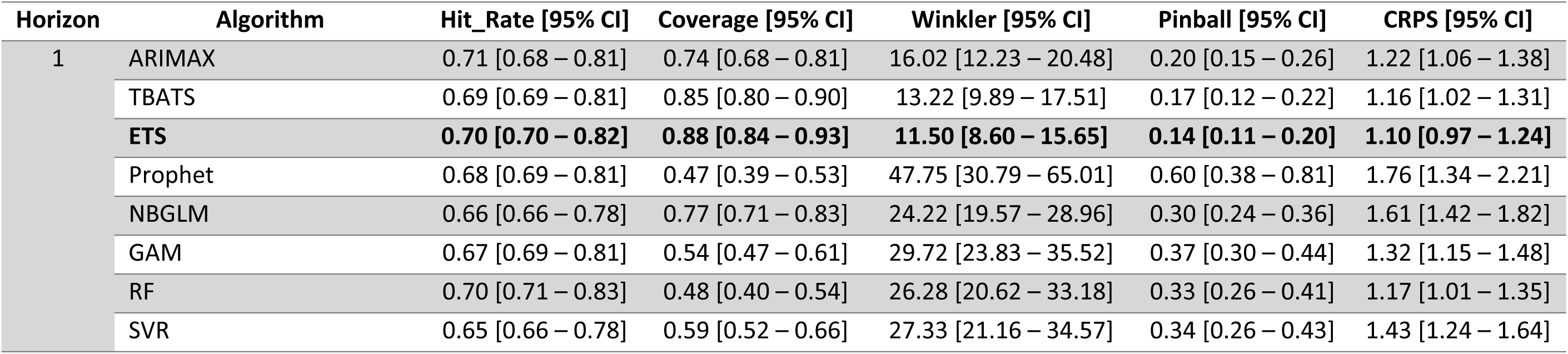
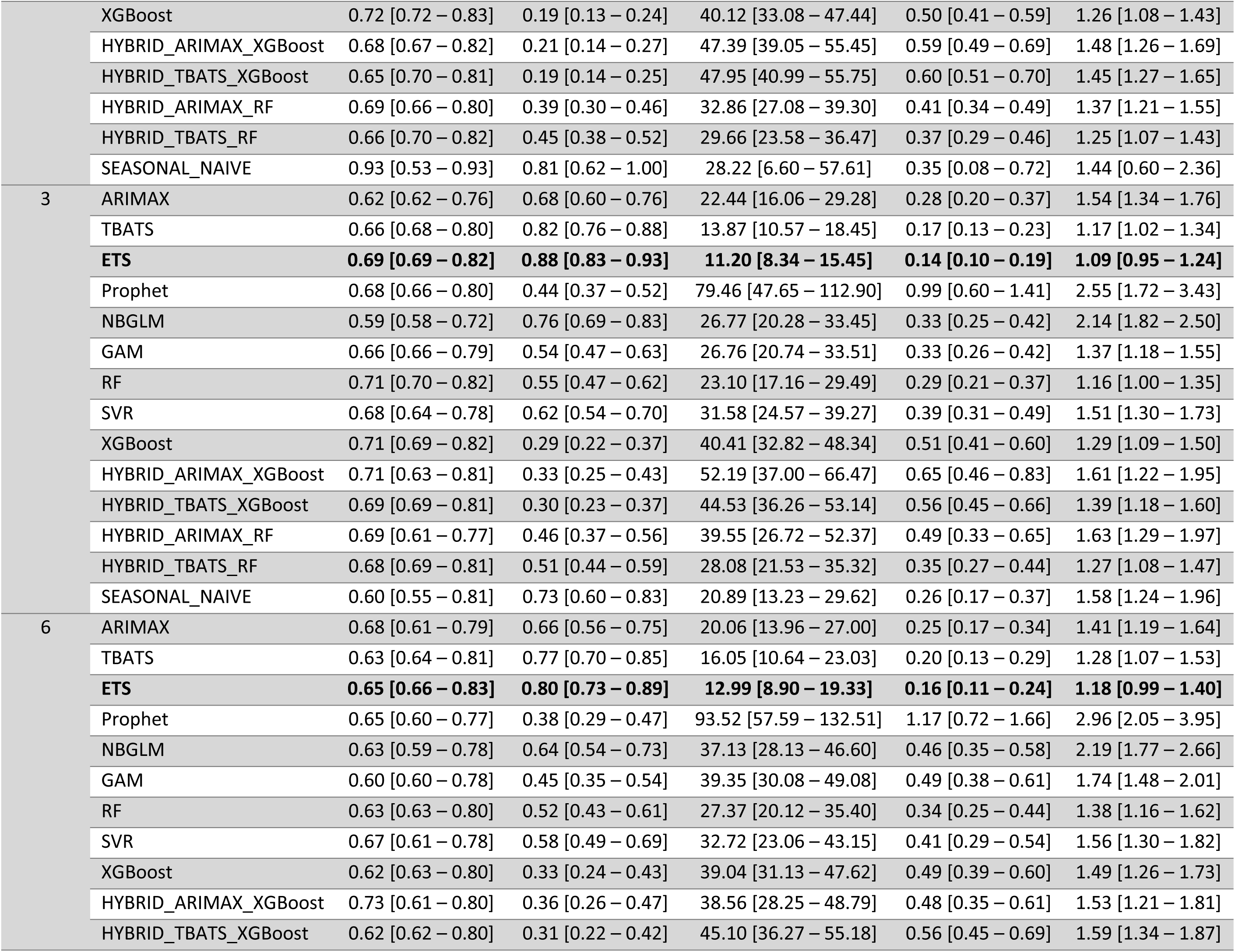
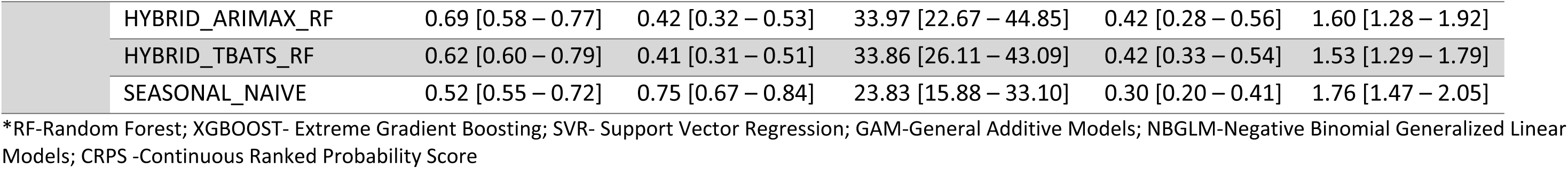
Model calibration and probabilistic metrics across horizons in *Shigella* forecasting.

| Horizon | Algorithm | Hit_Rate [95% CI] | Coverage [95% CI] | Winkler [95% CI] | Pinball [95% CI] | CRPS [95% CI] |
| --- | --- | --- | --- | --- | --- | --- |
| 1 | ARIMAX | 0.71 [0.68 – 0.81] | 0.74 [0.68 – 0.81] | 16.02 [12.23 – 20.48] | 0.20 [0.15 – 0.26] | 1.22 [1.06 – 1.38] |
|  | TBATS | 0.69 [0.69 – 0.81] | 0.85 [0.80 – 0.90] | 13.22 [9.89 – 17.51] | 0.17 [0.12 – 0.22] | 1.16 [1.02 – 1.31] |
|  | **ETS** | **0.70 [0.70 – 0.82]** | **0.88 [0.84 – 0.93]** | **11.50 [8.60 – 15.65]** | **0.14 [0.11 – 0.20]** | **1.10 [0.97 – 1.24]** |
|  | Prophet | 0.68 [0.69 – 0.81] | 0.47 [0.39 – 0.53] | 47.75 [30.79 – 65.01] | 0.60 [0.38 – 0.81] | 1.76 [1.34 – 2.21] |
|  | NBGLM | 0.66 [0.66 – 0.78] | 0.77 [0.71 – 0.83] | 24.22 [19.57 – 28.96] | 0.30 [0.24 – 0.36] | 1.61 [1.42 – 1.82] |
|  | GAM | 0.67 [0.69 – 0.81] | 0.54 [0.47 – 0.61] | 29.72 [23.83 – 35.52] | 0.37 [0.30 – 0.44] | 1.32 [1.15 – 1.48] |
|  | RF | 0.70 [0.71 – 0.83] | 0.48 [0.40 – 0.54] | 26.28 [20.62 – 33.18] | 0.33 [0.26 – 0.41] | 1.17 [1.01 – 1.35] |
|  | SVR | 0.65 [0.66 – 0.78] | 0.59 [0.52 – 0.66] | 27.33 [21.16 – 34.57] | 0.34 [0.26 – 0.43] | 1.43 [1.24 – 1.64] |
|  | XGBoost | 0.72 [0.72 – 0.83] | 0.19 [0.13 – 0.24] | 40.12 [33.08 – 47.44] | 0.50 [0.41 – 0.59] | 1.26 [1.08 – 1.43] |
|  | HYBRID_ARIMAX_XGBoost | 0.68 [0.67 – 0.82] | 0.21 [0.14 – 0.27] | 47.39 [39.05 – 55.45] | 0.59 [0.49 – 0.69] | 1.48 [1.26 – 1.69] |
|  | HYBRID_TBATS_XGBoost | 0.65 [0.70 – 0.81] | 0.19 [0.14 – 0.25] | 47.95 [40.99 – 55.75] | 0.60 [0.51 – 0.70] | 1.45 [1.27 – 1.65] |
|  | HYBRID_ARIMAX_RF | 0.69 [0.66 – 0.80] | 0.39 [0.30 – 0.46] | 32.86 [27.08 – 39.30] | 0.41 [0.34 – 0.49] | 1.37 [1.21 – 1.55] |
|  | HYBRID_TBATS_RF | 0.66 [0.70 – 0.82] | 0.45 [0.38 – 0.52] | 29.66 [23.58 – 36.47] | 0.37 [0.29 – 0.46] | 1.25 [1.07 – 1.43] |
|  | SEASONAL_NAIVE | 0.93 [0.53 – 0.93] | 0.81 [0.62 – 1.00] | 28.22 [6.60 – 57.61] | 0.35 [0.08 – 0.72] | 1.44 [0.60 – 2.36] |
| 3 | ARIMAX | 0.62 [0.62 – 0.76] | 0.68 [0.60 – 0.76] | 22.44 [16.06 – 29.28] | 0.28 [0.20 – 0.37] | 1.54 [1.34 – 1.76] |
|  | TBATS | 0.66 [0.68 – 0.80] | 0.82 [0.76 – 0.88] | 13.87 [10.57 – 18.45] | 0.17 [0.13 – 0.23] | 1.17 [1.02 – 1.34] |
|  | **ETS** | **0.69 [0.69 – 0.82]** | **0.88 [0.83 – 0.93]** | **11.20 [8.34 – 15.45]** | **0.14 [0.10 – 0.19]** | **1.09 [0.95 – 1.24]** |
|  | Prophet | 0.68 [0.66 – 0.80] | 0.44 [0.37 – 0.52] | 79.46 [47.65 – 112.90] | 0.99 [0.60 – 1.41] | 2.55 [1.72 – 3.43] |
|  | NBGLM | 0.59 [0.58 – 0.72] | 0.76 [0.69 – 0.83] | 26.77 [20.28 – 33.45] | 0.33 [0.25 – 0.42] | 2.14 [1.82 – 2.50] |
|  | GAM | 0.66 [0.66 – 0.79] | 0.54 [0.47 – 0.63] | 26.76 [20.74 – 33.51] | 0.33 [0.26 – 0.42] | 1.37 [1.18 – 1.55] |
|  | RF | 0.71 [0.70 – 0.82] | 0.55 [0.47 – 0.62] | 23.10 [17.16 – 29.49] | 0.29 [0.21 – 0.37] | 1.16 [1.00 – 1.35] |
|  | SVR | 0.68 [0.64 – 0.78] | 0.62 [0.54 – 0.70] | 31.58 [24.57 – 39.27] | 0.39 [0.31 – 0.49] | 1.51 [1.30 – 1.73] |
|  | XGBoost | 0.71 [0.69 – 0.82] | 0.29 [0.22 – 0.37] | 40.41 [32.82 – 48.34] | 0.51 [0.41 – 0.60] | 1.29 [1.09 – 1.50] |
|  | HYBRID_ARIMAX_XGBoost | 0.71 [0.63 – 0.81] | 0.33 [0.25 – 0.43] | 52.19 [37.00 – 66.47] | 0.65 [0.46 – 0.83] | 1.61 [1.22 – 1.95] |
|  | HYBRID_TBATS_XGBoost | 0.69 [0.69 – 0.81] | 0.30 [0.23 – 0.37] | 44.53 [36.26 – 53.14] | 0.56 [0.45 – 0.66] | 1.39 [1.18 – 1.60] |
|  | HYBRID_ARIMAX_RF | 0.69 [0.61 – 0.77] | 0.46 [0.37 – 0.56] | 39.55 [26.72 – 52.37] | 0.49 [0.33 – 0.65] | 1.63 [1.29 – 1.97] |
|  | HYBRID_TBATS_RF | 0.68 [0.69 – 0.81] | 0.51 [0.44 – 0.59] | 28.08 [21.53 – 35.32] | 0.35 [0.27 – 0.44] | 1.27 [1.08 – 1.47] |
|  | SEASONAL_NAIVE | 0.60 [0.55 – 0.81] | 0.73 [0.60 – 0.83] | 20.89 [13.23 – 29.62] | 0.26 [0.17 – 0.37] | 1.58 [1.24 – 1.96] |
| 6 | ARIMAX | 0.68 [0.61 – 0.79] | 0.66 [0.56 – 0.75] | 20.06 [13.96 – 27.00] | 0.25 [0.17 – 0.34] | 1.41 [1.19 – 1.64] |
|  | TBATS | 0.63 [0.64 – 0.81] | 0.77 [0.70 – 0.85] | 16.05 [10.64 – 23.03] | 0.20 [0.13 – 0.29] | 1.28 [1.07 – 1.53] |
|  | **ETS** | **0.65 [0.66 – 0.83]** | **0.80 [0.73 – 0.89]** | **12.99 [8.90 – 19.33]** | **0.16 [0.11 – 0.24]** | **1.18 [0.99 – 1.40]** |
|  | Prophet | 0.65 [0.60 – 0.77] | 0.38 [0.29 – 0.47] | 93.52 [57.59 – 132.51] | 1.17 [0.72 – 1.66] | 2.96 [2.05 – 3.95] |
|  | NBGLM | 0.63 [0.59 – 0.78] | 0.64 [0.54 – 0.73] | 37.13 [28.13 – 46.60] | 0.46 [0.35 – 0.58] | 2.19 [1.77 – 2.66] |
|  | GAM | 0.60 [0.60 – 0.78] | 0.45 [0.35 – 0.54] | 39.35 [30.08 – 49.08] | 0.49 [0.38 – 0.61] | 1.74 [1.48 – 2.01] |
|  | RF | 0.63 [0.63 – 0.80] | 0.52 [0.43 – 0.61] | 27.37 [20.12 – 35.40] | 0.34 [0.25 – 0.44] | 1.38 [1.16 – 1.62] |
|  | SVR | 0.67 [0.61 – 0.78] | 0.58 [0.49 – 0.69] | 32.72 [23.06 – 43.15] | 0.41 [0.29 – 0.54] | 1.56 [1.30 – 1.82] |
|  | XGBoost | 0.62 [0.63 – 0.80] | 0.33 [0.24 – 0.43] | 39.04 [31.13 – 47.62] | 0.49 [0.39 – 0.60] | 1.49 [1.26 – 1.73] |
|  | HYBRID_ARIMAX_XGBoost | 0.73 [0.61 – 0.80] | 0.36 [0.26 – 0.47] | 38.56 [28.25 – 48.79] | 0.48 [0.35 – 0.61] | 1.53 [1.21 – 1.81] |
|  | HYBRID_TBATS_XGBoost | 0.62 [0.62 – 0.80] | 0.31 [0.22 – 0.42] | 45.10 [36.27 – 55.18] | 0.56 [0.45 – 0.69] | 1.59 [1.34 – 1.87] |

|  |  |  |  |  |  |
| --- | --- | --- | --- | --- | --- |
| HYBRID_ARIMAX_RF | 0.69 [0.58 – 0.77] | 0.42 [0.32 – 0.53] | 33.97 [22.67 – 44.85] | 0.42 [0.28 – 0.56] | 1.60 [1.28 – 1.92] |
| HYBRID_TBATS_RF | 0.62 [0.60 – 0.79] | 0.41 [0.31 – 0.51] | 33.86 [26.11 – 43.09] | 0.42 [0.33 – 0.54] | 1.53 [1.29 – 1.79] |
| SEASONAL_NAIVE | 0.52 [0.55 – 0.72] | 0.75 [0.67 – 0.84] | 23.83 [15.88 – 33.10] | 0.30 [0.20 – 0.41] | 1.76 [1.47 – 2.05] |

\*RF-Random Forest; XGBOOST- Extreme Gradient Boosting; SVR- Support Vector Regression; GAM-General Additive Models; NBGLM-Negative Binomial Generalized Linear Models; CRPS -Continuous Ranked Probability Score

### Statistical Model Comparison

The Model Confidence Set (MCS) analysis revealed horizon-dependent dynamics in model distinguishability (Table 3). At the 1-month horizon, most models (12 of 14) were retained in the MCS, while NBGLM and SVR were excluded, indicating that these models performed significantly worse than the superior model set at α = 0.10. At the 3-month horizon, the MCS became more restrictive, with ARIMAX, NBGLM, GAM, SVR, HYBRID_ARIMAX_RF and SEASONAL_NAIVE excluded, while the remaining models were retained. At the 6-month horizon, most models were again retained, although SEASONAL_NAIVE was excluded. The pairwise comparison of the models based on DM are shown in are shown S1 Table. The observed and forecasts estimates for the best algorithms across each horizon (horizon 1=XGB, horizon 3= HYBRID_XGB_RF; horizon 6=ETS) are shown in Fig 3.

**Fig 3.**
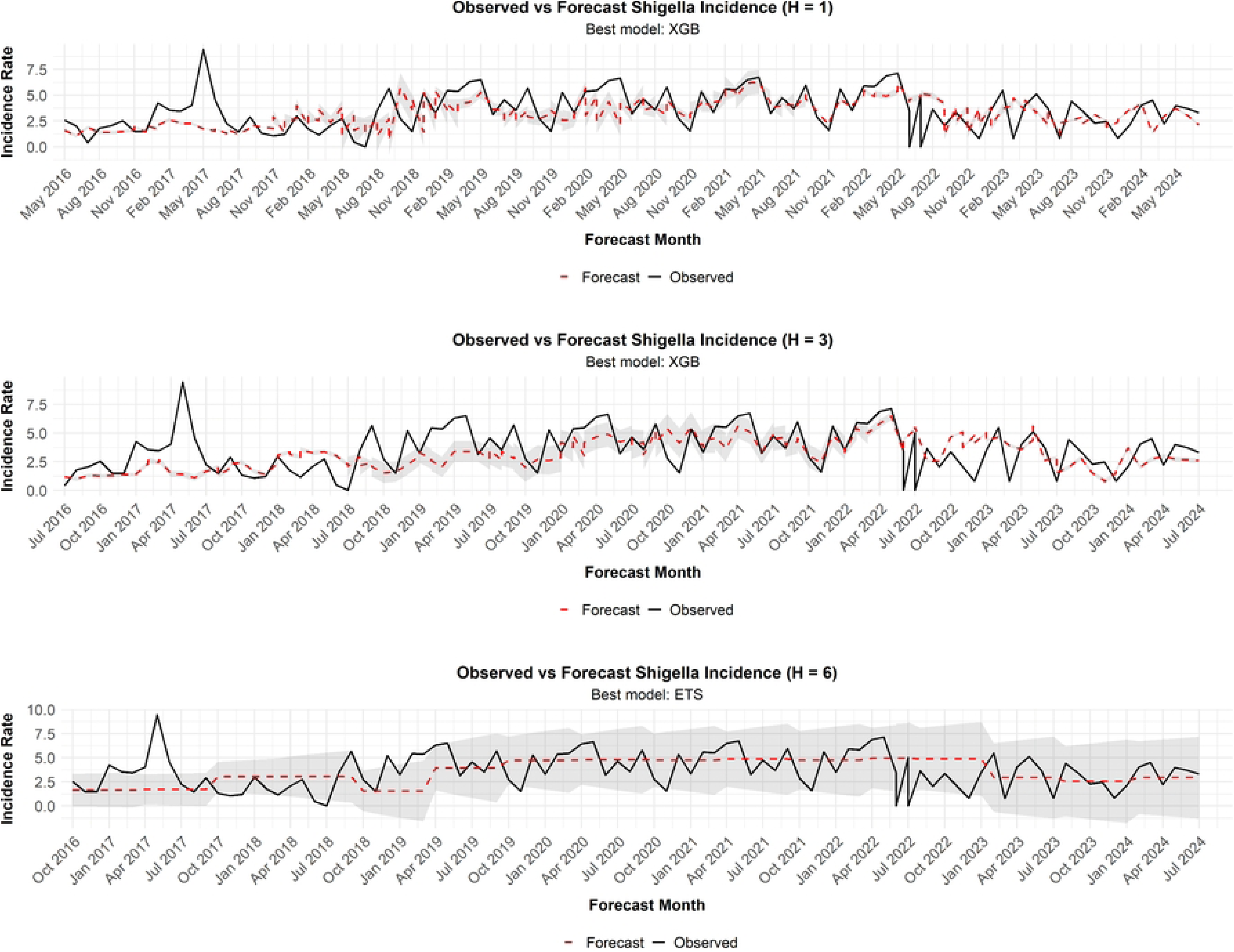
Observed and forecasted *Shigella* incidence across 1-, 3-, and 6-month forecasting horizons using the best-performing models, Siaya County, Kenya, 2008–2024

**Table 3.** Overall model comparison across horizons using model confidence set (MCS) in *Shigella* forecasting.

| Model | Horizon= 1 month |  |  | Horizon= 3 months |  |  | Horizon= 6 months |  |  |
| --- | --- | --- | --- | --- | --- | --- | --- | --- | --- |
|  | In MCS | Statistic | Alpha Level | In MCS | Statistic | Alpha Level | In MCS | Statistic | Alpha Level |
| ARIMAX | 1 | TR | 0.1 | 0 | TR | 0.1 | 1 | TR | 0.1 |
| TBATS | 1 | TR | 0.1 | 1 | TR | 0.1 | 1 | TR | 0.1 |
| ETS | 1 | TR | 0.1 | 1 | TR | 0.1 | 1 | TR | 0.1 |
| Prophet | 1 | TR | 0.1 | 1 | TR | 0.1 | 1 | TR | 0.1 |
| NBGLM | 0 | TR | 0.1 | 0 | TR | 0.1 | 1 | TR | 0.1 |
| GAM | 1 | TR | 0.1 | 0 | TR | 0.1 | 1 | TR | 0.1 |
| RF | 1 | TR | 0.1 | 1 | TR | 0.1 | 1 | TR | 0.1 |
| SVR | 0 | TR | 0.1 | 0 | TR | 0.1 | 1 | TR | 0.1 |
| XGBOOST | 1 | TR | 0.1 | 1 | TR | 0.1 | 1 | TR | 0.1 |
| HYBRID_ARIMAX_XGBOOST | 1 | TR | 0.1 | 1 | TR | 0.1 | 1 | TR | 0.1 |
| HYBRID_TBATS_XGBOOST | 1 | TR | 0.1 | 1 | TR | 0.1 | 1 | TR | 0.1 |
| HYBRID_ARIMAX_RF | 1 | TR | 0.1 | 0 | TR | 0.1 | 1 | TR | 0.1 |
| HYBRID_TBATS_RF | 1 | TR | 0.1 | 1 | TR | 0.1 | 0 | TR | 0.1 |
| SEASONAL_NAIVE | 1 | TR | 0.1 | 0 | TR | 0.1 | 1 | TR | 0.1 |

\*RF-Random Forest; XGBOOST- Extreme Gradient Boosting; SVR- Support Vector Regression; GAM-General Additive Models; NBGLM- Negative Binomial Generalized Linear Models; TR statistic- Range statistic
\* XGBoost- Extreme Gradient Boosting; ETS- statistical Exponential Smoothing

### Computational Efficiency

Computational efficiency varied across forecasting algorithms and horizons, spanning several orders of magnitude (Table 4). The seasonal-naive, NBGLM, ETS, and GAM models were the most computationally efficient, with runtimes <1 second per run (0.03–0.05, 0.14–0.20, 0.19–0.23, and 0.50–0.68 seconds, respectively) across horizons. Most other models required ∼ 8–80 seconds per run, including TBATS (11.55–12.89), ARIMAX (11.22–22.34), SVR (20.55–25.17), RF (39.24–50.22), and the hybrid models (7.63–30.88 seconds). Prophet was more variable (19.29–80.00 seconds). XGBoost was by far the most computationally intensive (1,041.31–1,079.03 seconds), despite its strong short-horizon point-forecast performance. Runtime showed no consistent relationship with forecast horizon but varied primarily by model architecture and optimization complexity.

**Table 4.** Computational efficiency of algorithms in *Shigella* forecasting across horizons.

| Algorithm | Horizon= 1 month | Horizon= 3 months | Horizon= 6 months |
| --- | --- | --- | --- |
|  | Execution in Seconds | Execution in Seconds | Execution in Seconds |
| ARIMAX | 14.06 | 22.34 | 11.22 |
| TBATS | 11.68 | 12.89 | 11.55 |
| ETS | 0.22 | 0.23 | 0.19 |
| Prophet | 19.42 | 80.00 | 19.29 |
| NBGLM | 0.20 | 0.15 | 0.14 |
| GAM | 0.68 | 0.68 | 0.50 |
| RF | 50.22 | 42.64 | 39.24 |
| XGB | 1052.77 | 1079.03 | 1041.31 |
| SVR | 23.47 | 20.55 | 25.17 |
| HYBRID_ARIMAX_RF | 30.88 | 18.77 | 17.59 |
| HYBRID_ARIMAX_XGB | 9.51 | 7.73 | 7.63 |
| HYBRID_TBATS_RF | 10.11 | 12.16 | 12.61 |
| HYBRID_TBATS_XGB | 19.43 | 18.55 | 18.53 |
| SEASONAL_NAIVE | 0.03 | 0.05 | 0.04 |

\*RF-Random Forest; XGBOOST- Extreme Gradient Boosting; SVR- Support Vector Regression; GAM-General Additive Models; NBGLM-Negative Binomial Generalized Linear Models;

### Cross-Horizon SHAP Evolution

SHAP analysis, conducted for the top performing model in each horizon, revealed a clear horizon-dependent shift in predictor importance, from environmental and seasonal forcing at short horizons to syndromic and healthcare utilization signals at longer horizons (Fig 4). At the 1-month horizon, predictions were dominated by secular temporal structure, seasonality, immediate climate conditions, and recent incidence. The linear trend was the strongest contributor, with higher values producing SHAP impacts of ≤ +1.2, followed by month and harmonic sine transforms (sin12), which captured intra-annual seasonal variation. Unlagged rainfall ranked fourth, with higher precipitation generally shifting predictions downward (SHAP ∼ −0.3), while unlagged temperature, 1-month incidence lag, and malaria burden amplified positive incidence predictions (≤ +0.5).

**Fig 4.**
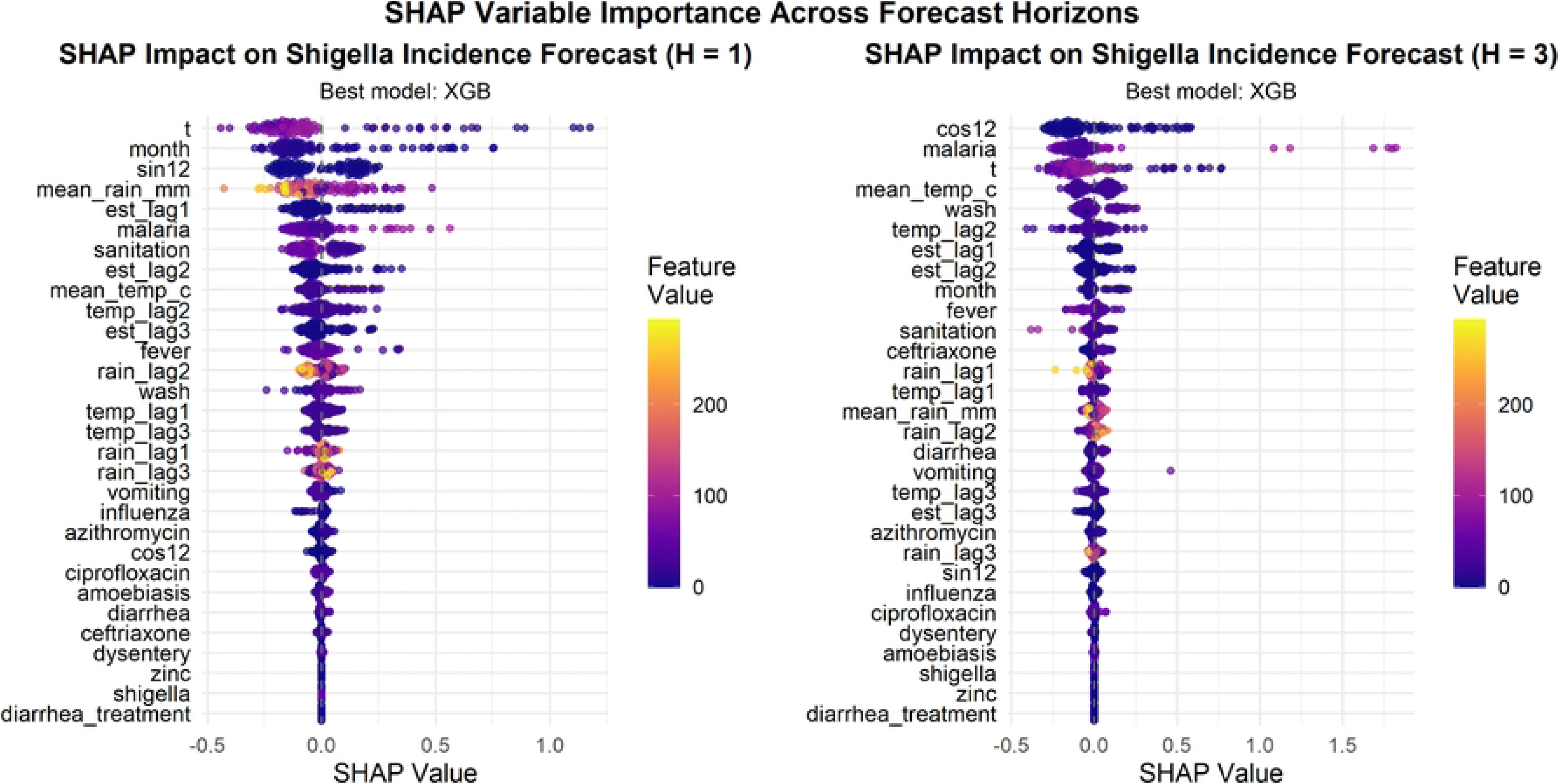
Horizon-specific SHAP attribution among best algorithms in *Shigella* forecasting, 2008-2024.

### * XGBoost- Extreme Gradient Boosting

At the 3-month horizon, attribution shifted toward annual cyclical structure, co-endemic disease dynamics, and environmental conditions. Cosine harmonic transform (cos12) emerged as the dominant predictor, capturing broader annual cycle timing. Malaria search indicators rose to the second most influential driver, producing substantial positive SHAP spikes (≥ +1.5) that demonstrate strong co-epidemiological risk amplification. Concurrent temperature and WASH also ranked among the leading predictors, driving elevated risk projections during high thermal periods. These findings indicate increasing reliance on broader seasonal and epidemiological patterns as the forecast horizon extended. Across both horizons, individual pharmaceutical usage indicators (diarrhea treatment, zinc, ceftriaxone) and disease search queries (*Shigella*) clustered around zero SHAP value, showing their limited predictive contribution. This confirms that macro-environmental, seasonal, and syndromic proxies provide far superior signal density for predicting community-level *Shigella* dynamics.

SHAP analysis was not conducted at the 6-month horizon because ETS was the best-performing algorithm at this horizon and, unlike the tree-based models, does not provide SHAP-compatible predictor attribution.

## Discussion

We evaluated several forecasting approaches, to determine models capable of providing accurate and operationally useful predictions for *Shigella* incidence in rural western Kenya, over a 16-year period, with the aim of informing early warning and public health preparedness. *Shigella* burden was substantial and heterogeneous across cohorts, with a mean incidence of 2.51 per 100 child-years. This multi-horizon forecasting study demonstrates that pediatric *Shigella* dynamics in Kenya can be forecast beyond seasonal persistence alone for up to six months. However, optimal model architectures and predictor sets shift systematically across forecasting horizons. XGBoost provided the strongest short-term point forecasts, whereas ETS offered the best longer-horizon performance and consistently superior uncertainty calibration. The higher point-prediction accuracy of ML models came at an important operational cost: XGBoost required >1,000 seconds per run and produced poorly calibrated prediction intervals, whereas ETS and TBATS provided well-calibrated forecasts with substantially lower computational requirements. Hybrid models consistently failed to outperform their single-model constituents. Although forecast errors increased with horizon, the best-performing models outperformed the seasonal-naïve benchmark, demonstrating incremental predictive value beyond seasonal persistence. Predictor importance similarly evolved from autoregressive and immediate environmental signals at 1-month to syndromic and pharmaceutical proxies at 3-months, and macro-seasonal and lagged climatic patterns at 6-months. Collectively, these findings support horizon-specific model selection that prioritizes not only point-forecast accuracy, but also probabilistic calibration and computational feasibility for operational early warning systems for shigellosis.

Our estimates of *Shigella* incidence across cohorts including KHIS data align with estimates from similar studies in the study setting [2,22,45]. *Shigella* remains a major cause of diarrheal morbidity, reinforcing the need for effective prevention strategies, including vaccination, to reduce both disease burden and downstream consequences such as antimicrobial use and resistance. The heterogeneity in incidence across cohorts reflects a changing epidemiological landscape [46] and highlights the importance of accounting for secular trends and differences in data sources when developing forecasts. Furthermore, periods bridged using routine KHIS data showed a slightly higher mean incidence, suggesting that routine health information systems can capture substantial endemic disease burden. This supports their use as a feasible complement to clinical trial and surveillance cohorts for sustaining longitudinal surveillance, particularly in resource-limited settings where primary data collection is constrained by limited resources and reductions in research funding [47].

Across all models, absolute errors increased with forecast horizon, reflecting the growing uncertainty inherent in multi-month infectious disease prediction. This observation is consistent with existing evidence [48], with some studies suggesting that performance degradation may be more pronounced for certain pathogens and peak transmission seasons. Nevertheless, the best-performing model at six months (ETS) reduced RMSE and MAE by approximately one-third relative to the seasonal-naïve benchmark, demonstrating that meaningful predictive skill persisted beyond the short term. This finding is consistent with a study from India forecasting acute diarrhea and dengue, which similarly reported superior performance of statistical, mathematical, and deep-learning models compared with benchmark models [49]. No single model dominated across horizons: XGBoost provided the strongest point forecasts at 1 and 3 months whereas ETS was superior at 6 months. This horizon-dependent performance supports adaptive model selection rather than reliance on a single forecasting algorithm. Tree-based machine-learning models therefore offered an advantage at shorter horizons, where recent and nonlinear signals were more informative, while ETS performed better at longer leads as predictive uncertainty increased and broader trend-seasonal structure became more important. Despite evidence from previous studies that hybrid approaches can combine nonlinear predictive power with improved calibration, our hybrid models did not consistently outperform their constituent algorithms, while more complex approaches such as Prophet and NBGLM also showed suboptimal performance. These findings are similar to those reported for meteorological forecasting by Mimi et al [50] and environmental modelling by Patel et al. [51]. This finding may be explained in part by the possibility that the constituent models already captured much of the predictable signal, particularly given that secular trend and autoregressive memory were among the most important predictors. Additionally, the value of the ML component may diminish as the forecast horizon increases, as longer-term forecasts become progressively less informed by recent temporal patterns and are subject to greater uncertainty. If it demonstrates that greater model complexity does not necessarily improve forecasting accuracy; models should therefore be selected based on demonstrable gains in predictive performance, calibration, and computational efficiency rather than architectural complexity alone.

We also observed a clear trade-off between point-forecast accuracy and probabilistic calibration consistent with findings from Camporeale and Carè [52]. Although XGBoost achieved the lowest point-forecast errors at the 1- and 3-month horizons, its prediction intervals were substantially under-covered, indicating overconfident uncertainty estimates. This suggests that, while tree-based ensembles can effectively capture nonlinear relationships in the conditional mean, deterministic implementations may not adequately represent predictive uncertainty in time-series settings without explicit probabilistic calibration. In contrast, ETS had slightly higher point-forecast errors at shorter horizons but consistently provided substantially better-calibrated prediction intervals. For public health applications, where forecasts inform risk stratification, preparedness, and resource allocation, reliable characterization of uncertainty is essential. Thus, model selection should consider both point-forecast accuracy and empirical calibration rather than relying on RMSE or MAE alone. At longer horizons, this distinction becomes even more important because ETS not only provided better-calibrated forecasts but also surpassed XGBoost in point-forecast performance at six months. These performance differences were accompanied by substantial variation in computational cost. XGBoost required approximately 1,000 seconds per run, compared with less than 1 second for ETS, NBGLM, GAM, and the seasonal-naïve model. Given that the advantage of XGBoost over ETS narrowed at three months and reversed at six months, its substantial computational burden may offer limited operational benefit, particularly for systems requiring frequent re-forecasting across multiple sites. In resource-constrained public health settings, parsimonious models such as ETS may therefore offer a more practical balance of accuracy, uncertainty calibration, and computational efficiency. More complex machine-learning and hybrid approaches should be favored only when their additional computational cost yields demonstrable improvements in predictive performance or calibration.

Predictor importance evolved systematically across forecast horizons, from autoregressive and immediate environmental signals at 1 month to syndromic and pharmaceutical proxies at 3 months, and broader seasonal and lagged climatic patterns at 6 months. This observation directly mirrors the multi-tiered architecture recommended by the WHO for integrated Early Warning Systems (EWS) validating the operational continuum bridging the gap between reactive, threshold-based outbreak detection used for immediate response and proactive seasonal environmental forecasting required for macroscopic resource positioning and structural epidemic preparedness [53]. At 1 month, predictions were driven primarily by recent incidence and concurrent temperature, reflecting short-term epidemic persistence and immediate environmental variation. This finding is congruent with observations from diarrheal forecasting in GEMS by Hossain et al. [54] and in China by Fang et al [55]. Autoregression captures epidemic persistence because a high volume of active shedding cases fuels rapid human-to-human transmission and heavily contaminates local environments for the following month. In parallel, high ambient temperatures drive immediate biological and behavioral shifts by causing rapid exponential multiplication of bacterial pathogens in water sources while instantly increasing human consumption of unboiled water. At 3 months, importance shifted toward diarrhea and azithromycin search volumes. These digital indicators may capture changes in community gastrointestinal illness and healthcare-seeking behavior that precede or accompany changes in *Shigella* activity, providing complementary information for medium-term forecasting. This finding is consistent with those in the forecasting of COVID-19 epidemic waves in Italy [56], and diarrheal related ED visits in South Korea [57]. By 6 months, the contribution of short-term autoregressive lags had diminished, with forecasts relying more heavily on broad seasonal cycles, lagged temperature, and malaria indicators. This horizon-dependent hierarchy indicates that *Shigella* forecasting operates across nested temporal scales, with local epidemic persistence dominating short-term forecasts and broader seasonal and epidemiological signals becoming increasingly informative at longer lead times. These findings support horizon-specific predictor selection rather than assuming constant predictor importance across forecast windows. Importantly, SHAP associations represent predictive signals rather than causal relationships to avoid designing spurious public health interventions.

Linear trend (*t*) was the strongest predictor across all horizons, complemented by lagged incidence, temperature, syndromic diarrhea, and azithromycin search volume. The dominance of the linear trend (t) across forecast horizons indicates that secular changes in *Shigella* incidence provided the most persistent source of predictive information, likely reflecting gradual shifts in population, disease ecology, and environmental conditions. Lagged incidence provided complementary autoregressive memory, particularly for short-term forecasts. This attenuation suggests that recent incidence becomes progressively less informative at longer lead times, with forecasts increasingly relying on broader seasonal and environmental signals. Temperature showed a similar horizon-dependent pattern, with concurrent temperature more informative at 1 month and lagged temperature gaining importance at 6 months, suggesting that delayed climatic signals may contribute to longer-range forecasting.

We also observed a co-endemicity with malaria where malaria search volumes emerged as a positive predictor at longer horizons, however this should not be interpreted as direct causality. Given that both *Shigella* and malaria peak during specific rainy and warm seasons in endemic regions, this finding likely captures shared environmental and seasonal risk windows and overlapping pediatric febrile and enteric surge dynamics rather than direct causation [58]. Conversely, we observed limited predictive utility from rainfall metrics, *Shigella* search volume, and specific individual therapeutics search volume (zinc, ceftriaxone, ciprofloxacin), proving that macro-seasonal and syndromic indicators carry much higher signal-to-noise ratios. This finding implies that healthcare utilization data and broad clinical syndromes can serve as potent leading indicators for culture-confirmed *Shigella* outbreaks, effectively capturing the epidemiological signal earlier than environmental drivers or internet search volumes (which had near-zero SHAP importance). This supports the integration of real-time pharmaceutical supply-chain data into early warning systems.

Our findings have important implications for both public health practice and forecasting methodology. The central implication is that effective *Shigella* forecasting requires balancing predictive accuracy, uncertainty calibration, computational efficiency, and information availability across forecast horizons. From a public health perspective, this supports integrating horizon-specific models into routine early-warning systems: short-term ML forecasts can inform tactical preparedness, including clinic-level outbreak staging and rehydration-supply preparation, whereas well-calibrated state-space models such as ETS/TBATS may be better suited to longer-term planning, including pre-emptive antimicrobial procurement, given their superior uncertainty calibration and computational efficiency. Furthermore, forecasts should communicate uncertainty explicitly, as overconfident predictions can mislead risk-based decisions. Additionally, the predictive contribution of diarrhea and azithromycin search volume, together with the feasibility of using KHIS data to bridge surveillance gaps, further supports integrating routinely collected syndromic and healthcare-utilization data with laboratory surveillance, particularly in resource-constrained settings where continuous intensive surveillance is difficult to sustain. However, SHAP-derived associations, including those involving malaria, should be interpreted as predictive signals rather than causal relationships and alone should not guide disease-specific interventions. Ultimately, forecasting can strengthen preparedness, but long-term *Shigella* control requires prevention. The persistent secular increase in incidence reinforces the need for *Shigella* vaccine introduction and sustained investment in WASH infrastructure to address the underlying burden beyond what short-term forecasting can achieve.

From a modelling perspective, the changing predictor hierarchy across horizons supports dynamic predictor selection and horizon-specific model development rather than static predictor sets. Model selection should jointly consider point accuracy, probabilistic calibration, computational cost, scalability, and ease of updating, with complex or hybrid approaches retained only when they provide demonstrable incremental value. The substantial computational burden of XGBoost relative to ETS highlights the importance of scalability for automated, multi-site forecasting across numerous sub-counties. Explicitly accounting for secular trends and non-stationarity will also be essential as the epidemiological trajectory evolves. Finally, SHAP and other attribution methods should be used to identify predictive signals rather than infer mechanisms, while environmental and digital-trace variables should be retained only when they demonstrate reproducible predictive benefit.

### Strengths and Limitations

This study has several strengths. First, it leveraged 16 years of monthly *Shigella* incidence data spanning multiple complementary surveillance platforms, utilizing standardized case definitions and weighting procedures to maximize comparability across study periods. Second, the nested rolling-origin evaluation scheme provided a realistic assessment of prospective forecasting capacity while strictly preventing temporal data leakage. Third, horizon-specific dynamic predictor selection and hyperparameter tuning allowed predictor importance and model architectures to adapt flexibly to varying lead times. Fourth, evaluating classical statistical, machine learning, and hybrid models against a baseline seasonal-naïve benchmark across point accuracy, probabilistic calibration, inferential robustness, and computational runtimes ensured a comprehensive operational benchmarking framework. Finally, integrating SHAP attribution enhanced model interpretability by transparently mapping predictor contributions across forecast horizons.

Despite these strengths, some limitations warrant consideration. First, gaps in the continuous incidence series required state-space Kalman smoothing and state-space imputation, introducing uncertainty tied to assumptions of temporal stability between active surveillance and routine health-system data. Second, inherent differences in study design, case ascertainment, and data aggregation across underlying surveillance platforms (GEMS, VIDA, EFGH, and KHIS) may have left residual measurement heterogeneity despite harmonization and weighting. Third, Google Trends search volumes and spatially aggregated meteorological metrics serve as indirect proxies that may not fully capture hyper-local transmission dynamics or healthcare-seeking behavior. Finally, while SHAP predictor attributions clarify model decision dynamics, these associations remain strictly predictive rather than causal; consequently, model performance and predictor importance may not generalize directly beyond rural western Kenya or to settings with contrasting epidemiological and health-seeking profiles.

### Conclusion

In rural western Kenya, *Shigella* incidence was forecastable beyond seasonal persistence alone for up to six months, but the sources and reliability of predictive information changed systematically with forecast horizon. Short-term forecasts benefited most from recent incidence, temporal structure, and immediate environmental conditions, whereas longer-horizon forecasts increasingly relied on broader seasonal, syndromic, climatic, and co-epidemiological signals. XGBoost provided superior short-term point forecasts, while ETS offered the best longer-horizon performance and balance of accuracy, uncertainty calibration, and computational efficiency, demonstrating that superior point accuracy does not necessarily translate into reliable probabilistic forecasts. These findings support horizon-specific, dynamically informed early-warning systems that integrate routinely available surveillance and syndromic data and evaluate both predictive accuracy and probabilistic reliability. However, forecasting can strengthen preparedness but cannot replace prevention; the persistent secular increase in *Shigella* burden reinforces the need for vaccination and sustained WASH investment as essential long-term control strategies.

## Materials and Methods

### Ethical Considerations

The protocol for the current study was reviewed and approved by the Scientific and Ethical Review Unit (SERU) of the Kenya Medical Research Institute (KEMRI) with the approval granted under KEMRI SERU#5416. Additionally, the protocols for GEMS, VIDA, and EFGH studies were reviewed and approved by the SERU with the approvals granted under KEMRI SSC Protocol #1155, SERU#2996 and SERU#4362, respectively. Moreover, GEMS and VIDA obtained approval from the Institutional Review Board (IRB) of the University of Maryland, School of Medicine with approvals granted under UMB Protocol #H-28327 for GEMS and UMB Protocol #: HM-HP-00062472 for VIDA. Furthermore, the IRB for the Centers for Disease Control and Prevention, Atlanta, GA, USA approved the VIDA protocol (reliance agreement 6729) and formally deferred its review of the GEMS protocol to the University of Maryland IRB (CDC Protocol # 5038). Prior to initiating any study procedures, written informed consent was obtained from each caregiver of all participating children across all three studies.

### Study Population and Data Sources

We estimated monthly *Shigella* incidence among children under five years in Kenya using primary individual-level data from two large case-control studies and a hybrid surveillance study: the Global Enteric Multicenter Study (GEMS) conducted from 2008 to 2011, the Vaccine Impact on Diarrhea in Africa (VIDA) study carried out from 2015 to 2018 and EFGH (August 2022 – July 2024). The underlying designs and methods for GEMS, VIDA, and EFGH have been previously described [17–20]. GEMS and VIDA were case-control studies that used similar methodologies to recruit children under five years of age seeking care for moderate-to-severe diarrhea (MSD). In contrast, EFGH enrolled all children aged 6–35 months presenting with diarrhea. To ensure cross-study standardization, we restricted the EFGH data to children meeting the GEMS/VIDA MSD criteria (diarrhea with dehydration, dysentery, or hospitalization). To resolve the inter-study data gaps (2014/01-2015/06 and 2018/08-2022/07), a syndromic-to-pathogen extrapolation framework was implemented using the KHIS [21]. Data from these primary sources were accessed on 15/05/2026 for the present analysis.

### Estimation of Monthly Incidence GEMS, VIDA, EFGH and KHIS

Incidence estimates for GEMS, VIDA, and EFGH were derived using a three-stage weighting approach that accounted for (i) case and healthy under-five (from Healthcare Utilization Survey) control sampling, (ii) healthcare-seeking behavior, and (iii) pathogen-specific positivity rates, as described elsewhere [2,22]. Child-years of observation were obtained from the Kenya Medical Research Institute Health and Demographic Surveillance System (HDSS), with monthly population estimates available from 2008–2024 [12,23]. Healthcare-seeking weights were estimated using monthly logistic regression models fitted to HUS controls, incorporating age, sex, diarrhea duration, stool frequency, dysentery, vomiting, fever, and their two-way interactions. Each case was weighted by the inverse predicted probability of seeking care at a study facility, with additional weights applied for care-seeking at study sentinel facilities out of those who sought care at any facility.

For *Shigella*, pathogen-specific weighting was based on the inverse of the monthly proportion of participants testing positive for *Shigella* such that the cases selected for testing were representative of the overall enrolled population and adequately reflected seasonal variation in diarrheal etiology. The monthly weighted number of medically attended *Shigella* cases was calculated as the sum of enrollment, healthcare-seeking, stool-testing, and pathogen-specific weights applied to *Shigella*-positive cases. Monthly incidence (per 100 child-years) was estimated by dividing the weighted number of cases by the corresponding child-years of observation.

Uncertainty was quantified using a dual-frame bootstrap with 1,000 replicates. HUS participants were resampled with replacement from the full control pool, while cases were resampled independently within each calendar month, and the entire weighting and incidence estimation procedure was repeated. Ninety-five percent confidence intervals were derived from the bootstrap distribution using log-scale Wald intervals. For months with fewer than five non-missing HUS partcipants, the log-scale standard error was borrowed from the pooled standard deviation across months with complete bootstrap data.

Aggregate monthly *Shigella*-positive cases during the missing periods were imputed by applying an enrollment cap alongside the average monthly quantitative PCR *Shigella* positivity rates derived from the active surveillance cohorts to the total monthly diarrheal case counts among children under five years reported in KHIS. The enrolment cap was implemented to ensure balanced seasonal and geographic representation of enteric pathogens during the same period and for KHIS we implemented the aggregated monthly enrolment cap. For these months, incidence was estimated using aggregated monthly estimates of *Shigella*-positive cases derived from KHIS as described above. To account for under-ascertainment, we applied a seasonal inflation factor derived from GEMS and VIDA data, calculated as the average ratio of weighted to observed *Shigella*-positive cases for each calendar month. For each calendar month, the correction factor was based on the ratio of the weighted number of *Shigella*-positive cases to the corresponding observed number of *Shigella*-positive cases. This correction accounted for differential healthcare seeking and case ascertainment. The adjusted number of medically attended cases was obtained by multiplying the estimated *Shigella*-positive count by the corresponding monthly inflation factor, and incidence was calculated per 100 child-years using monthly population estimates. When a month-specific inflation factor was unavailable, the overall mean inflation factor was used.

Additionally, missing values in the incidence series, where KHIS data were not available (2011/02-2011/10; 2012/10-2013/12), were imputed using a covariate-informed state-space model implemented with the KFAS package in R. Specifically, a local linear trend model was specified with monthly mean temperature and total rainfall (both scaled to z-scores) as external regressors. Unknown variances of the trend, observation error, and regression coefficients were estimated via maximum likelihood (BFGS), and the Kalman smoother was applied to reconstruct the full incidence series.

### Covariate Data Extraction

The forecasting incorporated a comprehensive set of health, healthcare-seeking and treatment-related digital indicators, climatic and environmental covariates, and temporal and seasonal terms to capture potential determinants of the observed trends. For 15 health-and behavior-related indicators (diarrhea, dysentery, *Shigella*, amoebasis, azithromycin, ciprofloxacin, ceftriaxone, sanitation, zinc, diarrhea treatment, fever, malaria, vomiting, Influenza and WASH), we obtained Google Trends (GT; Alphabet Inc, Mountain View CA, USA) relative search interest data for Kenya from January 2008 through December 2024 [24]. Where available, we used Google Trends topics (curated concepts) to minimize language-related bias; otherwise, the corresponding text-based search term was used.

Monthly mean 2-m air temperature was obtained from ERA5 reanalysis (Copernicus Climate Data Store) for the Siaya County bounding box (0.25°N, 33.90°E to −0.10°S, 34.45°E), converted from Kelvin to Celsius, and spatially averaged to a single monthly value [25]. Monthly rainfall was derived from CHIRPS (0.05° resolution) by extracting global monthly data, cropping to the same bounding box, and computing the spatial mean [26]. Both climate series were subsequently lagged by 1, 2, and 3 months for analysis.

A linear time trend (t) and seasonal Fourier terms (sin12 = sin(2πt/12), cos12 = cos(2πt/12)) were also added. The final dataset comprised a balanced monthly time series covering January 2008 to July 2024.

### Forecasting Framework

The primary evaluation used a rolling-origin design [27] with an initial 96-month (8-year) training window, a 12-month completely held-out evaluation period, and a 6-month step between successive forecast origins. At each origin, models were trained only on observations available before the forecast origin and evaluated on the subsequent held-out period. Forecasts were generated at 1-, 3-, and 6-month horizons. For each horizon, the training data were transformed into a direct forecasting problem by pairing predictors observed at time *t* with incidence at time *t+h*. This procedure produced horizon-specific supervised datasets and ensured that predictor selection and model tuning were conducted separately for each forecast horizon. The schematic overview of our *Shigella* forecasting pipeline is shown in Fig 5.

**Fig 5.**
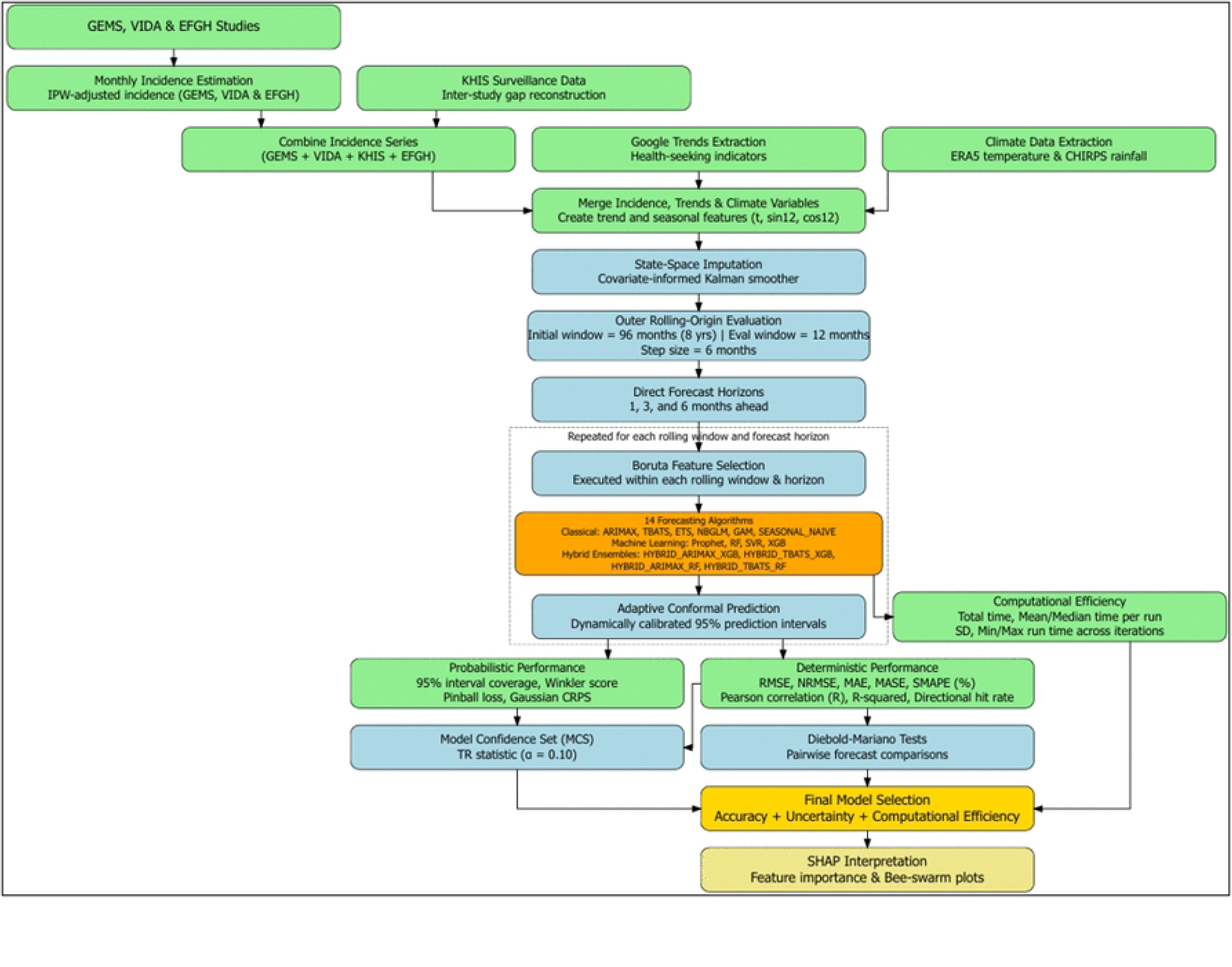
Schematic overview of the *Shigella* forecasting pipeline.

Hyperparameters for RF, SVR, XGBoost, and hybrid ML components were optimized within each rolling training window using blocked temporal cross-validation with expanding training sets and six-month validation blocks. The hyperparameter combination with the lowest mean validation RMSE was selected and refitted to the full training window. For random forest, the tuning grid varied the number of candidate variables at each split (mtry), minimum terminal node size (5, 10, or 20), and number of trees (300 or 500). For support vector regression, a radial basis function kernel was used, and the cost parameter (C), kernel parameter (gamma), and epsilon-insensitive loss parameter (epsilon) were varied over prespecified grids. For XGBoost, the tuning grid varied learning rate (eta), maximum tree depth, minimum child weight, and number of boosting rounds (nrounds), while subsampling and column subsampling were fixed at 0.8. The same temporal tuning strategy was applied to the ML components of the hybrid forecasting models.

### Predictor Selection with Boruta

For each outer rolling-origin window, Boruta (a random forest-based wrapper method) was applied to select relevant predictors [28]. Boruta compares the importance of original variables against that of shadow randomized copies, retaining variables that are significantly more important than random. The algorithm was run for up to 50 iterations (or until all variables were confirmed/rejected). Only the selected predictors were passed to the forecasting models, reducing dimensionality and mitigating overfitting. Variable importance scores (mean Z-score across iterations) were summarized across rolling windows using mean importance and selection frequency, allowing assessment of both predictive contribution and temporal stability.

### Forecasting Algorithms

The fourteen forecasting algorithms were organized into three complementary classes: classical statistical/state-space models, machine learning (ML) methods, and residual-learning hybrid architectures designed to combine their respective strengths. Classical models included ARIMAX, which extends autoregressive integrated moving average processes by incorporating exogenous covariates to capture linear temporal dependence and external drivers [29]; statistical Exponential Smoothing (ETS), a state-space exponential smoothing framework that adaptively models level, trend, and seasonality using weighted historical information [30]; TBATS, which accommodates complex and potentially multiple seasonal cycles through trigonometric representations, Box–Cox transformation, and ARMA error structures [31]; a Negative Binomial GLM, which models over-dispersed count outcomes via a log-link specification [32]; and Generalized Additive Model (GAM), which relaxes parametric assumptions by using smooth spline functions to capture nonlinear relationships [33].

The ML class comprised Random Forest (RF), an ensemble of decorrelated decision trees that captures high-order nonlinear interactions through bootstrap aggregation [34]; Extreme Gradient Boosting (XGBoost), a regularized gradient-boosting framework that sequentially optimizes prediction error using additive tree learners [35]; Support Vector Regression (SVR) with a radial basis function kernel, which maps predictors into a high-dimensional predictor space to learn nonlinear relationships within an ε-insensitive loss framework [36]; and Prophet, an additive decomposition model combining piecewise trends, Fourier-based seasonality, and covariate effects [37].

Finally, four hybrid residual-learning models were constructed by combining structured time-series models with flexible ML corrections. In these frameworks, ARIMAX or TBATS first captured global temporal structure and seasonality, after which RF or XGBoost was trained on the remaining residuals to learn nonlinear, local deviations, yielding a two-stage decomposition of signal and error. A seasonal-naïve model was included as the benchmark. The complete model set was evaluated separately at each forecast horizon.

### Prediction Intervals and Uncertainty Quantification

Prediction intervals were estimated using model-specific approaches: parametric uncertainty from classical models, residual bootstrap procedures for ML models, and Gaussian error propagation for nonlinear learners. To improve calibration, we applied an adaptive conformal prediction (ACP) scheme [38]. For each model and forecast horizon, a scaling parameter (γ, initialized at 0) was updated sequentially based on empirical coverage: γ was increased by 0.025 when observations fell outside the prediction interval and decreased by 0.05 when they fell inside. The final interval was obtained by scaling the nominal width as [point − width(1+γ), point + width(1+γ)], enabling horizon-specific, online recalibration that adaptively corrects under-or over-coverage across rolling windows.

### Evaluation Metrics and Model Comparison

Forecast performance was evaluated through a structured framework encompassing point accuracy, probabilistic calibration, and inferential robustness [39–41]. For deterministic point accuracy, we assessed both magnitude and directional fidelity using a comprehensive set of metrics. Root Mean Square Error (RMSE), normalized RMSE (NRMSE), and Mean Absolute Error (MAE), mean absolute scaled error (MASE), quantified absolute predictive error, capturing sensitivity to large deviations, while Symmetric Mean Absolute Percentage Error (sMAPE) provided a scale-independent measure of relative accuracy. Pearson correlation (R) and the coefficient of determination (R²) evaluated the strength of linear association and variance explained by the forecasts. Directional accuracy assessed the ability of models to correctly capture the sign of temporal changes, and calibration was examined through regression of observed on predicted values, summarized by slope and intercept to detect systematic bias.

Probabilistic performance focused on the quality of predictive uncertainty rather than point estimates alone. Empirical 95% interval coverage assessed calibration against nominal uncertainty levels, while the Winkler score jointly penalized interval width and mis-coverage to evaluate sharpness and reliability. Pinball loss was used to assess quantile accuracy across lower and upper bounds, and Continuous Ranked Probability Score (CRPS) provided an overall measure of probabilistic distance between predictive distributions and observed outcomes under a Gaussian approximation derived from interval widths.

Uncertainty quantification and model comparison were supported by rigorous inferential procedures. The Model Confidence Set (MCS) identified a subset of statistically indistinguishable best-performing models using a stepwise elimination approach with pairwise Diebold–Mariano (DM) tests used to evaluate differences in predictive accuracy across models while accounting for horizon-specific dependence [42,43]. To ensure robustness, all metrics were accompanied by 1000 bootstrap resamples of forecast– observation pairs, yielding non-parametric confidence intervals without distributional assumptions.

Computational efficiency was assessed by recording elapsed time for predictor selection, model tuning, and model fitting within each forecast horizon and algorithm. Runtime was retained as an operational performance characteristic alongside predictive accuracy and probabilistic calibration to assess the feasibility of deploying models in repeated and multi-site forecasting systems.

### SHapley Additive exPlanations (SHAP) Analysis for Interpretability

For the best-performing model at each forecast horizon, predictor contributions were interpreted using SHAP [44] implemented via the *fastshap* package. A model-specific prediction wrapper was defined for both tree-based models and linear models to enable consistent attribution. SHAP values were estimated using 30 Monte Carlo permutations per observation, with adjustments to account for predictor correlation structure. Results were summarized using beeswarm plots, where each point represents an individual SHAP value, colored by the corresponding predictor value, and predictors are ranked according to mean absolute SHAP importance. SHAP-derived associations were interpreted as predictive signals rather than causal effects.

All modelling and statistical analyses were carried out using R software, version 4.6.0 (R Foundation for Statistical Computing, Vienna, Austria). The following key packages were used: KFAS, forecast, prophet, Boruta, ranger, xgboost, e1071, mgcv, fastshap, ggplot2, MCS, and dplyr.

## Acknowledgements

We extend our deep gratitude to the families who participated in the three studies, and to the clinical and field teams whose dedication and hard work made this research possible. We are also grateful to the physicians, administrators, and Ministry of Health officials at each recruitment center in Siaya County for their provision of facilities, operational support, and gatekeeper permissions that enabled data access and the successful implementation of the studies.

## Data Availability

The GEMS and VIDA datasets are publicly available on ClinEpiDB (https://clinepidb.org/ce/app) while the EFGH data will is publicly available through the Vivli data-sharing platform at https://search.vivli.org/doiLanding/studies/PR00011860/isLanding.

## Supplemental Materials

S1 Fig. Boruta-derived predictor selection across forecasting horizons for *Shigella* incidence, 2008–2024.

S1 Table: Pairwise model comparison across horizons using Diebold-Mariano (DM) tests

